# The real-life impact of vaccination on COVID-19 mortality in Europe and Israel

**DOI:** 10.1101/2021.05.26.21257844

**Authors:** Katarzyna Jabłońska, Samuel Aballéa, Mondher Toumi

## Abstract

**OBJECTIVES:** This study aimed at estimating the real-life impact of vaccination on COVID-19 mortality, with adjustment for SARS-CoV-2 variants spread and other factors across Europe and Israel.

**METHODS:** Time series analysis of daily number of COVID-19 deaths was performed using non-linear Poisson mixed regression models. Variants’ frequency, demographic, climate, health and mobility characteristics of thirty-two countries were considered as potentially relevant adjustment factors between January 2020 and April 2021.

**RESULTS:** The analysis revealed that vaccination efficacy in terms of protection against deaths was equal to 72%, with a lower reduction of number of deaths for B.1.1.7 versus non-B.1.1.7 variants (70% and 78%, respectively). Other factors significantly related to mortality were arrivals at airports, mobility change from the pre-pandemic level and temperature.

**CONCLUSIONS:** Our study confirms a strong effectiveness of COVID-19 vaccination based on real-life public data, although lower than expected from clinical trials. This suggests the absence of indirect protection for non-vaccinated individuals. Results also show that vaccination effectiveness against mortality associated with the B.1.1.7 variant is slightly lower compared with other variants. Lastly, this analysis confirms the role of mobility reduction, within and between countries, as an effective way to reduce COVID-19 mortality and suggests the possibility of seasonal variations in COVID-19 incidence.

## Introduction

The pandemic of the coronavirus infectious disease 2019 (COVID-19) is continuously evolving, driven by the spread of new variants of severe acute respiratory syndrome coronavirus 2 (SARS-CoV-2). During the second half of 2020 and early 2021 variety of new SARS-CoV-2 variants emerged. EU2 variant (mutation S:447N) firstly observed in July 2020 in western Europe, was found to be capable of increasing virus infectivity.^1,2^ Then, several variants of concern (VOC) have been identified, including B.1.1.7 developed firstly in the UK in September 2020,^3^B.1.351 in South Africa in December 2020,^4^ P.1 in Brazil in January 2021,^5^ and the ‘Indian’ variant B.1.617 reported firstly in Maharashtra in January 2021.^6^ The disease mortality has been increased in these countries after new variants were developed.^7-10^ An increased risk of transmissibility, hospitalization and death associated with the B.1.1.7 variant was reported by number of authors.^8,11-16^ The B.1.351 variant was found to have an increased transmissibility and immune escape,^17^ and was estimated to be 50% more transmissible than preexisting variants.^18^ Higher incidence of COVID-19 cases in younger age groups were observed in the Amazonas state, suggesting changes in pathogenicity of the P.1 variant.^19^ Preliminary findings suggest also a significant increase in case fatality rate in young and middle-aged for the P.1 mutant.^20^ The region of Maharashtra, where the B.1.617 variant emerged, experienced significant rise in daily infection rate after the new variant appeared.^10^

To control the SARS-CoV-2 spread, numbers of different vaccines have been developed and analyzed in clinical trials, including eight vaccines having emergency use or conditional marketing authorizations worldwide or across regions, as of May 2021.^21^ The worldwide vaccination campaign has started in December 2020 aiming to provide herd immunity across societies. The threshold for COVID-19 “herd immunity” was placed between 60–70% of the population gaining immunity through vaccinations or past disease exposure, however, scientists warn that herd-immunity is unlikely to be achieved due to factors such as vaccine hesitancy and the spread of new variants.^22,23^ Israel was far ahead of other countries in terms of the proportion of vaccinated inhabitants, exceeding 62% at the end of April 2021, with the UK reaching 50% and the USA 42% at the same time.^24^

Results of clinical trials on vaccine efficacy revealed that Pfizer-BioNTech had 95% efficacy at preventing symptomatic COVID-19 infection in people without prior infection.^25^ 94.1% efficacy was reported for Moderna,^26^ 70.4% for Oxford-AstraZeneca,^27^ 66.5% for Johnson & Johnson^28^ and 96.4% for Novavax,^29^ with the latter being still under the investigation before authorization. For the prevention against a severe disease course, Pfizer, Moderna, AstraZeneca and Novavax reported a 100% efficacy, whereas 84% was observed for Johnson & Johnson, however, the latter was tested on a broader range of countries, including the USA, South Africa and Brazil, after the new VOCs spread.

Clinical evidence suggests that newly developed virus variants may affect the protective efficacy of both naturally acquired immunity and of vaccinations. Studies on neutralization of convalescent sera against distinct strains showed that VOCs were harder to neutralize than the original strain, an early Wuhan-related strain of SARS-CoV-2. Neutralization titers against the B.1.1.7 variant showed a 3-fold reduction,^30^ a 3.4-fold reduction was observed for the P.1 variant,^31^ and a 13.3-fold reduction for the B.1.531 variant.^32^ Johnson & Johnson vaccine was found to have 64% efficacy against infection in South Africa and 68% in Brazil after the spread of B.1.135 and P.1 variants, whereas the efficacy against severe-critical disease was 82% and 88% in both countries, respectively.^28^ Late March 2021, AstraZeneca was observed as having 70.4% efficacy against the B.1.1.7 variant,^33^ however, the vaccine did not protect as well against B.1.351 variant.^34^ For Novavax, the initial evidence suggests 86.3% efficacy against B.1.1.7 variant^35^ and 49.4% against B.1.351.^36^ Early May 2021, Pfizer vaccine was found to be 87% effective against infection with B.1.1.7 variant and 72% against B.1.351 variant, whereas 97.4% efficacy against severe disease course was observed for any of these mutations.^37^ On the other hand, Wang (et al.) observed a reduced neutralization of Moderna and Pfizer vaccine-immune sera against B.1.351 variant (12.4-fold for Moderna; 10.3-fold for Pfizer), but no significant impact was observed for B.1.1.7 variant.^38^ Next, a 3.8-4.8-fold reduced neutralization of Moderna vaccine was observed against P.1 variant.^31^ Preliminary evidence suggests a significant drop in neutralization of B.1.617 compared to other variants, including B.1.1.7 and P.1, with sera of Indian’s vaccine, Covaxine.^10^

The other concern is the probability of re-infection after recovery or vaccination. Hansen (et al.) observed an 80.5% protection against re-infection in a population-level observational study on Danish patients previously tested positive for SARS-CoV-2,^39^ however, the study was performed before VOCs spread. The probability of re-infection after vaccination is also a big concern. As reported by the USA Centers for Disease Control and Prevention (CDC), there were around 9200 infections among vaccinated inhabitants among 95 million of those who have already been vaccinated in the USA (0.01%) as of 26 April 2021.^40^ Despite this optimistic preliminary data, effects of vaccination were not reliably assessed in a longer perspective thus far. The real-world evidence on vaccination effectiveness is still sparse. Experts alarm that additional data are needed to assess the potential impact of VOCs on future vaccine efficacy.^41^ Considering all the concerns associated with new VOCs spread, the real vaccination effectiveness becomes hard to assess and prognose but can be expected to decrease over time. Also, it is likely that vaccination may favour the emergence of new variants by selection of new, better fitted mutants. Some scientists suggest that, similarly as for seasonal flu vaccines, COVID-19 vaccines will need to be redesigned or even updated periodically to protect against new variants.^42,43^

Vaccination efficacy and distinct variants spread are only two factors among numerous other variables affecting COVID-19 infection and death rates across the world. A variety of potential predictors were assessed in the literature, including demographic characteristics, mobility and social-distancing measures, environmental and climate variables, as well as health characteristics.^44-53^

This study aims at estimating the real-life impact of vaccination on COVID-19 mortality based on publicly available data from Europe and Israel, using time series analysis with non-linear mixed regression models. Variants frequency, including B.1.1.7 and other variants, as well as country-specific demographic and meteorological characteristics, health indicators and mobility factors were considered as potentially relevant adjustment factors. Results of the current study should inform policy decision makers, scientists, and the general public about the role of vaccination and social-distancing strategies in controlling the COVID-19 pandemic in the face of new VOCs spread.

## Methods

### Data collection

A total of 32 countries were considered in the analysis, including European countries and Israel. The daily number of COVID-19 deaths was the primary outcome of interest. Values were smoothed using 7-day moving average, divided by the number of inhabitants of a given country and reported as daily numbers of deaths per 1 million inhabitants.

The main explanatory variables of interest were proportion of vaccinated inhabitants (vaccination coverage), as well as average proportions of SARS-CoV-2 variants calculated across strains forming twelve Nextstrain clades. The focus was on 20A (EU2), 20E (EU1) and 20I (B.1.1.7) variants, with two formers being dominant in Europe during the summer 2020, and the latter VOC being most frequent early 2021. Other time-varying covariates considered in the analysis were maximum daily temperature, mean daily wind speed, number of arrivals at two biggest airports of a country and change in mobility from pre-pandemic level (considering the average across retail/recreation, transit stations and groceries/pharmacies). Additional fixed covariates were proportion of population aged 65 or more, prevalence of diabetes and rate of cardiovascular deaths.

Data on COVID-19 deaths and vaccination were obtained from Our World In Data on 15 April 2021.^24^ Meta-data on SARS-CoV-2 virus variants (clades) identified up to mid-April 2021 was downloaded from the Nextstrain platform.^54-56^ We assumed that if a strain was observed on a given date, it could be observed in a range of ±14 days from the observation date. Since the data were not reported daily, linear interpolation was used to impute missing observations, assuming zeros a month before the first and after the last (if up to 1 March 2021) reported occurrence of a variant. Finally, data were smoothed with the use of 14-day moving average.

Countries’ characteristics were obtained from Our World In Data, Eurostat, National Centers for Environmental Information, Aviation Intelligence Portal and Google COVID-19 Community Mobility Reports.^24,57-60^ Data on arrival flights and mobility were smoothed using 7-day moving average.

### Statistical analysis

Regression analysis was used to investigate the association between COVID-19 mortality and daily reported time-varying variables and fixed covariates.

The primary analysis of daily number of COVID-19 deaths was performed with the use of non-linear Poisson mixed model with random country-level intercept and mobility effects. The considered period was from the date of first reported death in Europe, 29 January 2020, up to 15 April 2021.

Due to the presence of autocorrelations, and to consider the fact that the number of infections on a given day is dependent on the number of infectious cases in the population over previous days, the model was adjusted for the logarithm of daily number of COVID-19 deaths reported 7 days earlier. To capture the fact that increasing or decreasing trends in COVID-19 mortality over time are generally stable over several weeks or months, the logarithm of quotient of COVID-19 deaths 7 days before divided by deaths 14 days before the actual date was added as a covariate. All other time-varying variables were considered with a 21-day lag, to account for the virus incubation period and the delay between being infected and die due to the disease. In addition, heterogeneity between countries was considered with random intercepts and mobility effects varying between countries.

Assuming *M* indicates mortality with a vaccination coverage *‘c’, M*_*O*_ is the mortality without vaccination and *‘VE’* represents the vaccine efficacy, we have:

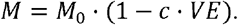

After applying the logarithmic transformation and considering a set of covariates *x*_1_,…,*x*_*k*_ and random effects *u*_0_,*u*_1_,…,*u*_*n*_ on intercept and selected *x*_1_,…,*x*_*n*_, the equation above was extended as shown below to specify the non-linear model:

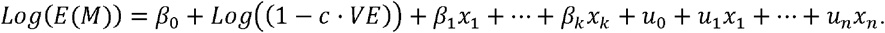

For the exploratory analysis, vaccine efficacy against B.1.1.7 and non-B.1.1.7 variants was analyzed using a similar approach. Assuming that there are two classes of virus variants with known proportions equaled *p*_1_ and *p*_2_, the vaccine efficacy could be considered as the average efficacy weighted by variants proportions:

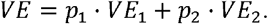

The formula for the non-linear model is then as follows:

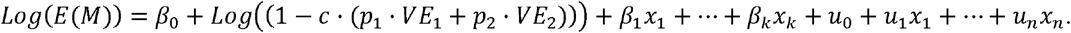

A p-value lower than 0.05 was considered as statistically significant. Akaike’s Information Criterion (AIC) was provided to inform about models’ fit statistic. Analyses were performed using SAS 9.4 software.

## Results

### Descriptive statistics

Descriptive statistics of outcomes and covariates across 32 countries included in the analysis, for the period between January 2020 and April 2021 are presented in Table 1. Cumulative numbers of COVID-19 deaths per inhabitant across countries during this period are presented in Figure 1. The highest numbers were observed for central and south-east countries: Czechia, Hungary, Bosnia and Herzegovina. Mean proportions of SARS-CoV-2 variants, EU2, EU1 and B.1.1.7 for each country are presented in Figure 2. Up to mid-April 2021, the variant EU2 was the most frequently spread for vast majority of countries, except Israel and the UK for which B.1.1.7 was more frequent, as well as Spain and Lithuania with EU1 being more commonly observed.

**Table 1.** Descriptive statistics of outcomes and covariates across 32 countries (January 2020 – April 2021)

| Variable | N | Mean | Standard Deviation | Median | Lower Quartile | Upper Quartile | Minimum | Maximum |
| --- | --- | --- | --- | --- | --- | --- | --- | --- |
| Number of daily COVID-19 deaths | 13229 | 3.355 | 4.618 | 1.159 | 0.206 | 4.933 | 0 | 28.770 |
| Proportion of vaccinated inhabitants | 13181 | 0.018 | 0.058 | 0 | 0 | 0 | 0 | 0.616 |
| Proportion of 20A (EU2) variant | 13106 | 0.349 | 0.250 | 0.313 | 0.154 | 0.509 | 0 | 1.000 |
| Proportion of 20E (EU1) variant | 12754 | 0.090 | 0.159 | 0 | 0 | 0.114 | 0 | 0.741 |
| Proportion of 20I (B.1.1.7) variant | 13099 | 0.124 | 0.256 | 0 | 0 | 0.065 | 0 | 1.000 |
| Max daily temperature | 13008 | 15.597 | 9.347 | 15.833 | 8.750 | 22.389 | -17.111 | 43.111 |
| Mean daily wind speed | 13010 | 6.227 | 3.353 | 5.600 | 3.700 | 8.100 | 0.100 | 25.800 |
| Arrivals at two biggest airports | 12748 | 140.609 | 195.648 | 66.429 | 27.714 | 177.929 | 1.857 | 1780.143 |
| Mobility change from pre-pandemic<sup>1</sup> | 12918 | -0.217 | 0.170 | -0.201 | -0.322 | -0.091 | -0.792 | 0.199 |
| Proportion of inhabitants aged ≥65 | 13229 | 0.184 | 0.025 | 0.190 | 0.168 | 0.198 | 0.117 | 0.230 |
| Diabetes prevalence | 13229 | 0.062 | 0.019 | 0.058 | 0.048 | 0.072 | 0.033 | 0.101 |
| Cardiovascular death rate | 13229 | 2.037 | 1.124 | 1.535 | 1.148 | 2.783 | 0.861 | 5.398 |

<sup>1</sup> Average mobility change from pre-pandemic calculated across retail/recreation, transit stations and groceries/pharmacies [0.01]. Descriptive statistics were calculated for 32 countries based on daily data between 29 January 2020 and 15 April 2021.

**Figure 1.**
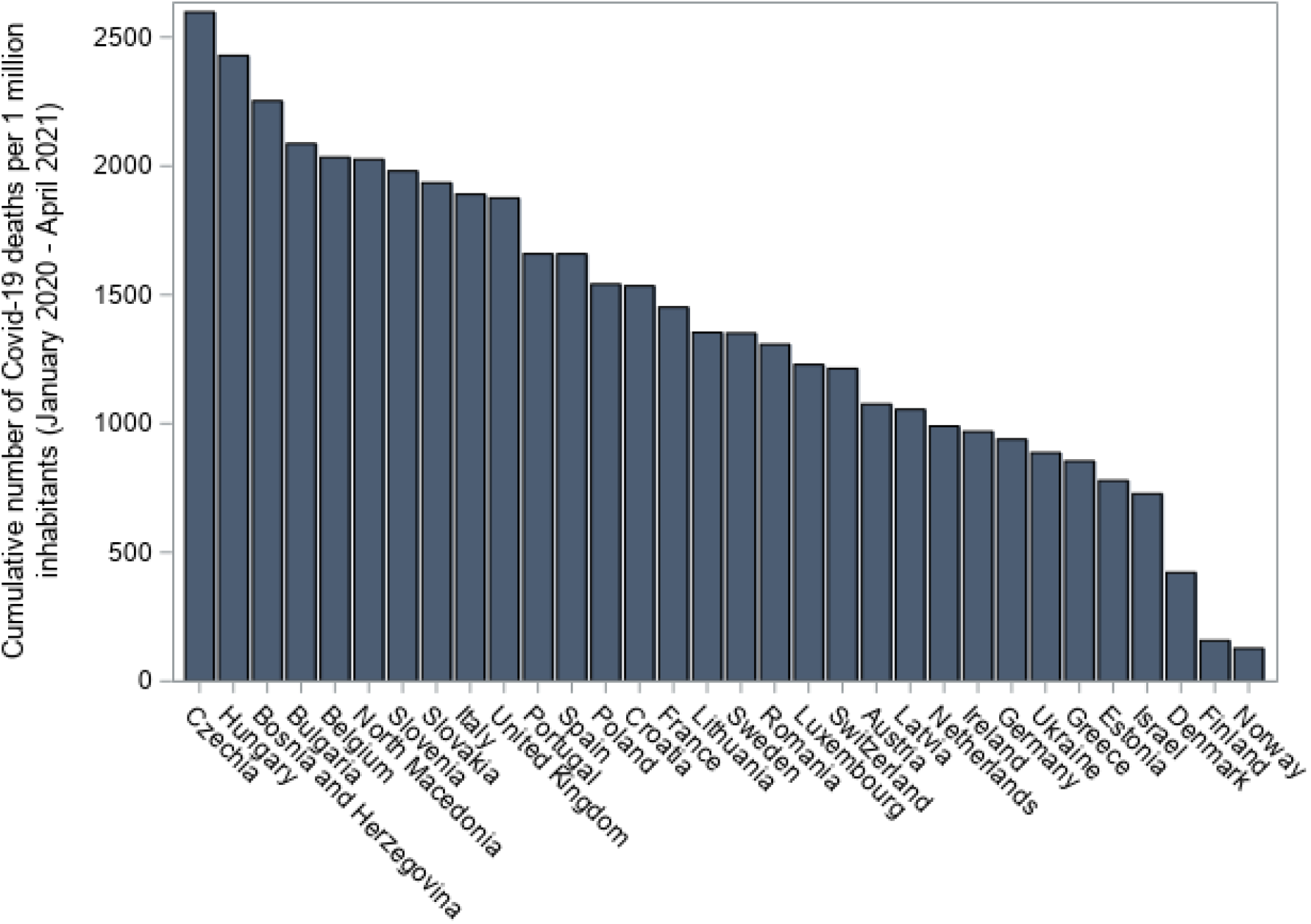
Cumulative number of COVID-19 deaths per 1 million inhabitants across countries. Cumulative number of COVID-19 deaths per one million inhabitants were calculated based on data on mortality between 29 January 2020 and 15 April 2021.

**Figure 2.**
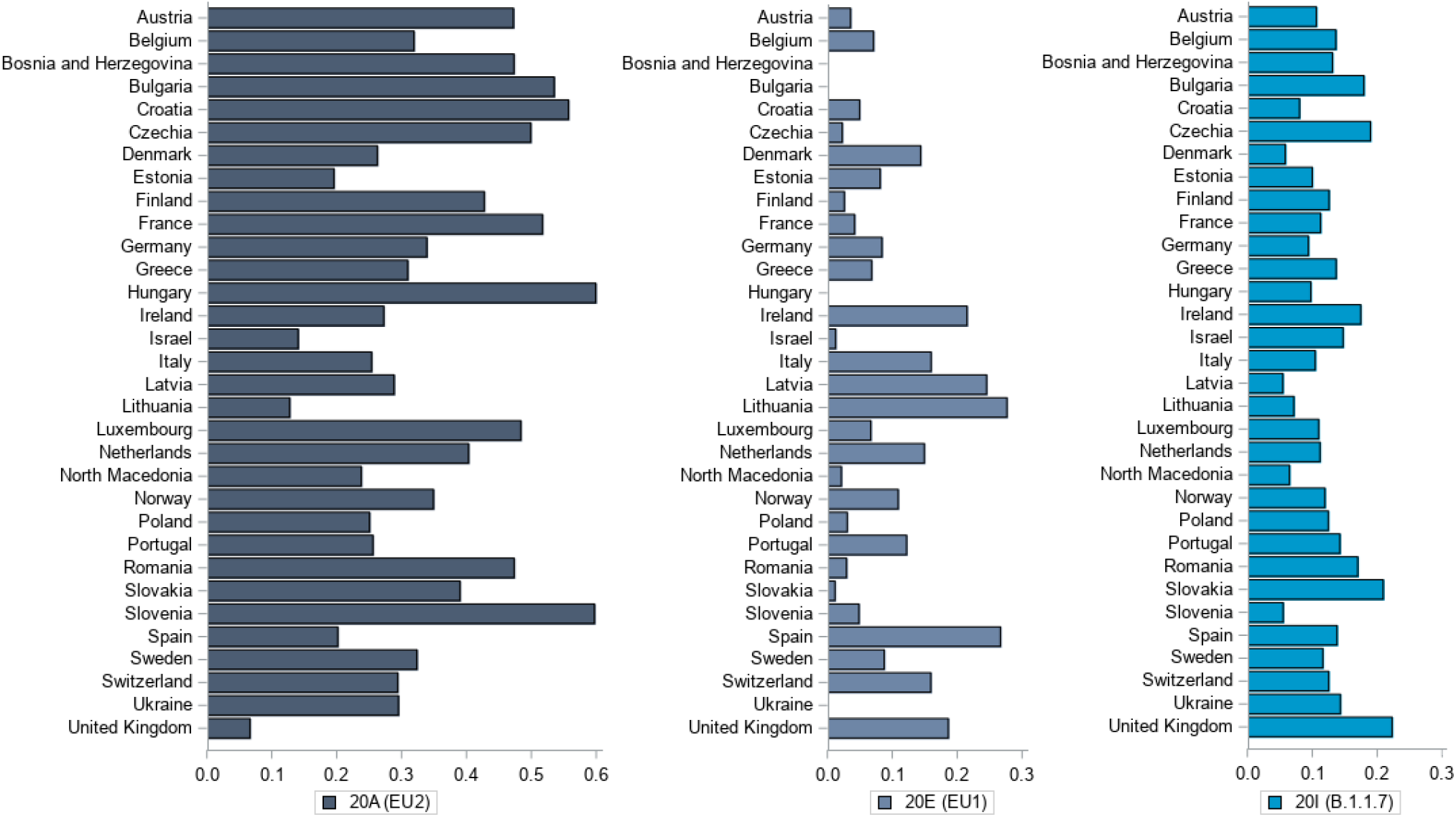
Mean SARS-CoV-2 variants proportions across countries, between January 2020 and April 2021. Mean proportions of variants were calculated based on daily data on variants proportions across observed strains used to form Nextstrain clades (variants), between 29 January 2020 and 15 April 2021.

### Primary analysis

Analysis of the non-linear Poisson mixed model of number of COVID-19 deaths revealed that the effect of vaccination efficacy against mortality was assessed as significant and equalled to 0.720 (p<0.001; Table 2). Other covariates which were found significant in the model were: temperature (−0.005, p<0.001), arrivals at airports (0.709, p<0.001) and mobility change from pre-pandemic level (0.753, p<0.001). Variables used to account for autocorrelation and minimize the effect of trend were assessed as significant (Log of number of daily COVID-19 deaths: 0.926, p<0.001; Log of number of COVID-19 deaths 7 days before / 14 days before: 0.158, p<0.010).

**Table 2.** Results of the non-linear Poisson mixed model of number of daily COVID-19 deaths.

| Variable | Estimate | Standard Error | P value |
| --- | --- | --- | --- |
| Intercept | 0.467 | 0.202 | 0.028 |
| Logarithm of number of daily COVID-19 deaths<sup>#</sup> | 0.926 | 0.007 | <0.001 |
| Logarithm of (number of COVID-19 deaths 7 days before / 14 days before) | 0.158 | 0.015 | <0.001 |
| Proportion of 20A (EU2) variant* | 0.012 | 0.036 | 0.746 |
| Proportion of 20E (EU1) variant* | -0.032 | 0.046 | 0.501 |
| Proportion of 20I (B.1.1.7) variant* | 0.059 | 0.033 | 0.084 |
| Max daily temperature* | -0.005 | 0.001 | <0.001 |
| Mean daily wind speed* | -0.002 | 0.002 | 0.351 |
| Arrivals at two biggest airports [in thousands]* | 0.709 | 0.085 | <0.001 |
| Mobility change from pre-pandemic<sup>*1</sup> | 0.753 | 0.064 | <0.001 |
| Proportion of inhabitants aged ≥65<sup>^</sup> | -1.238 | 0.930 | 0.193 |
| Diabetes prevalence<sup>^</sup> | 0.544 | 1.293 | 0.677 |
| Cardiovascular death rate<sup>^</sup> | 0.023 | 0.023 | 0.322 |
| Vaccination efficacy* | 0.720 | 0.132 | <0.001 |
| Variance for RE on intercept | 0.014 | 0.006 | 0.023 |
| Variance for RE on mobility | 0.036 | 0.023 | 0.126 |

<sup>#</sup>Time-varying variable with a 7-days lag.
\* Time-varying variable with a 21-days lag.
<sup>^</sup>Fixed variable.
<sup>1</sup> Average mobility change from pre-pandemic calculated across retail/recreation, transit stations and groceries/pharmacies [0.01].
Abbreviations: AIC = Akaike's Information Criterion; mln = million; RE = random effects.
Non-linear Poisson mixed model of daily number of COVID-19 deaths per 1 mln inhabitants, with country-specific random effects on intercept and mobility. Daily data for 32 countries were included, from 29 January 2020 up to 15 April 2021.

### Exploratory analysis

Results of the analysis of the exploratory model revealed numerically lower vaccine efficacy against B.1.1.7 than against non-B.1.1.7 variants, although the difference was not statistically significant (0.697, p=0.002 and 0.778, p=0.049, respectively; Table 3). The same set of covariates were found significant in the exploratory model as in the primary analysis: temperature (−0.005, p<0.001), arrivals at airports (0.703, p<0.001) and mobility change from pre-pandemic level (0.753, p<0.001). Variables used to account for autocorrelation and minimize the effect of trend were assessed as significant (Log of number of daily COVID-19 deaths: 0.926, p<0.001; Log of number of COVID-19 deaths 7 days before / 14 days before: 0.158, p<0.010).

**Table 3.** **Results of the non-linear Poisson mixed model of number of daily COVID-19 deaths with interactions between variants proportions and vaccination efficacy**

| Variable | Estimate | Standard Error | P value |
| --- | --- | --- | --- |
| Intercept | 0.409 | 0.204 | 0.054 |
| Logarithm of number of daily COVID-19 deaths<sup>#</sup> | 0.926 | 0.007 | <0.001 |
| Logarithm of (number of COVID-19 deaths 7 days before / 14 days before) | 0.158 | 0.015 | <0.001 |
| Proportion of 20A (EU2) variant* | 0.012 | 0.036 | 0.747 |
| Proportion of 20E (EU1) variant* | -0.034 | 0.047 | 0.467 |
| Proportion of 20I (B.1.1.7) variant* | 0.059 | 0.033 | 0.085 |
| Max daily temperature* | -0.005 | 0.001 | <0.001 |
| Mean daily wind speed* | -0.002 | 0.002 | 0.346 |
| Arrivals at two biggest airports [in thousands]* | 0.703 | 0.085 | <0.001 |
| Mobility change from pre-pandemic<sup>*1</sup> | 0.753 | 0.064 | <0.001 |

| **Variable** | **Estimate** | **Standard Error** | **P value** |
| --- | --- | --- | --- |
| Proportion of inhabitants aged $\geq 65^{\wedge}$ | -0.805 | 0.925 | 0.391 |
| Diabetes prevalence<sup>^</sup> | 0.124 | 1.285 | 0.924 |
| Cardiovascular death rate<sup>^</sup> | 0.025 | 0.023 | 0.278 |
| Vaccination efficacy against 20I (B.1.1.7)* | 0.697 | 0.201 | 0.002 |
| Vaccination efficacy against variants other than 20I (B.1.1.7)* | 0.778 | 0.379 | 0.049 |
| *Variance for RE on intercept* | 0.013 | 0.006 | 0.025 |
| *Variance for RE on mobility* | 0.035 | 0.023 | 0.132 |

AIC = 30186.
<sup>#</sup> Time-varying variable with a 7-days lag.
\* Time-varying variable with a 21-days lag.
<sup>^</sup> Fixed variable.
<sup>1</sup> Average mobility change from pre-pandemic calculated across retail/recreation, transit stations and groceries/pharmacies [0.01].
Abbreviations: AIC = Akaike's Information Criterion; mln = million; RE = random effects.
Non-linear Poisson mixed model of daily number of COVID-19 deaths per 1 mln inhabitants, with country-specific random effects on intercept and mobility, including interactions between variants proportions and vaccination efficacy. Daily data for 32 countries were included, from 29 January 2020 up to 15 April 2021.

## Discussion

In this study we investigated the association between daily mortality due to COVID-19 and vaccination coverage, proportions of SARS-CoV-2 variants and additional factors, such as demographics, health, mobility and meteorological variables, analyzing country-level data across Europe and Israel. Results of the analysis suggest that vaccination effectiveness against deaths is equal to 72%, and that it is slightly lower against the B.1.1.7 variant compared to non-B.1.1.7 variants (difference not statistically significant). These findings suggest lower effectiveness against death than reported efficacy against severe or critical disease course in clinical trials of vaccines (84-100%).^25-29^

This lower than expected effectiveness might be explained by the difference in considered populations: clinical trials included restrictive populations and our study covers general populations, irrespective of age, concomitant therapies, medical condition and general condition. In particular, vaccinated people in real life are older on average than subjects enrolled in clinical trials (12.2% aged ≥55 in AstraZeneca trial; 24.7% aged ≥65 in Moderna trial; 33.5% aged ≥60 in Johnson & Johnson trial; 42.3% aged ≥55 in Pfizer trial). However, our results suggesting lower protection against B.1.1.7 variant, are consistent with reported data so far from in vivo experiments and patient level studies, providing an external validation of these findings. Laboratory evidence revealed a slight reduction in neutralization against B.1.1.7 variant compared to the original strain. Neutralization titers against this VOC were 3-fold lower when analyzing convalescent sera, and 3.3-fold and 2.5-fold lower for Pfizer and AstraZeneca vaccinees, respectively.^30^ Real-world studies on B.1.1.7 VOC suggested that it caused increased mortality compared to non-B.1.1.7 variants,^15,61^ which, therefore, might not have been contained with similar effectiveness by vaccination. Our results evoke the question of variants evading vaccine antibodies in the future and the need to adapt such vaccine for each new season, which was earlier suggested by experts.^42,43^

While the utilization of individual-level data, collected in real-world setting, could provide more precise estimates of vaccine effectiveness, the use of aggregate data at country level also has a major advantage: the vaccination impact estimated in this analysis should capture the indirect protection provided by vaccination. If the vaccine protects against infection, the number of infectious cases would decrease as more people are vaccinated. The lower number of infectious cases in the population would lead to a reduced probability for susceptible individuals to get in contact with infectious cases, thus leading to a reduction in incidence among all people, including non-vaccinated people. This indirect protection can be captured when comparing different populations with different rates of vaccination coverage, but could not be captured when comparing vaccinated and non-vaccinated individuals from the same population. Interestingly, the fact that our estimated vaccine effectiveness is relatively low compared to vaccine efficacy reported in clinical trials suggests that there is no or little indirect protection provided by vaccination. This could indicate that the vaccine protects against disease but not against infection, or that vaccinated groups of population are not those that contribute to the propagation of the virus.

A positive relationship between number of arrivals at airports and mortality has been observed in this analysis, similarly as between mobility change and mortality. It suggests that both increased long-distance travel and increased mobility are strong predictors of growth in daily number of COVID-19 deaths. These findings highlight the role of mobility reduction, both within and between countries, as an effective way to reduce COVID-19 mortality, especially when new virus variants emerge and spread across the world. Our results are in line with previous study by Jabłońska (et al.) suggesting that countries with lower reduction in mobility at the beginning of the pandemic experienced a higher COVID-19 daily deaths peak.^50^ The role of social distancing was also underlined by Badr (et al.) who observed a significant impact of mobility on COVID-19 transmission in the USA.^62^

The daily temperature was found as a significant predictor of COVID-19 mortality in this study, with increasing temperature associated with the reduction in number of deaths. Kerr (et al.) found no consensus on the impact of meteorological factors on COVID-19 spread in their literature review, however, they suggested existence of environmental sensitivity of COVID-19, but not as significant as non-pharmaceutical interventions and human behavior.^63^ Several authors underlined that disease seasonality may exist,^64-67^ including Liu (et al.) who found that COVID-19 infection and mortality rates were higher in colder climates and that the cold season caused an increase in total infections, while the warm season contributed to the opposite effect.^66^ Since our analysis covered a full annual cycle of COVID-19, our result suggests the possibility of seasonal variations in COVID-19 incidence. Such seasonality has been well established in temperate climate for other respiratory viruses.^68,69^

### Limitations

Our study has several limitations. Firstly, our analysis was conducted on a country-level basis to estimate the vaccination efficacy, which should be seen as less precise method compared to analysis of individual-level data, as previously noted. However, given the range of included countries, our results shed light on the problem of vaccination effectiveness from a broader perspective and investigate the effect of vaccination across societies, considering variability of vaccination coverage through time and between countries. Secondly, the quality of data on variants distribution varied between countries and was low for some of them, therefore, results of the exploratory analysis should be treated with cautious. To limit bias and avoid fluctuations, we employed methods of interpolation and smoothing. Countries with limited data were excluded. Thirdly, the set of covariates used in the multivariate analysis can be assessed as non-exhaustive. We decided to consider factors which were previously assessed as significantly impacting the risk of severe illness or mortality from COVID-19 in the literature.^50-53^ Finally, we were not able to consider other new SARS-CoV-2 VOCs, except B.1.1.7, in the current analysis, which was due to their limited spread in Europe as of April 2021. It rises a need for further research on this topic in the future.

### Conclusions

This study confirms a strong effectiveness of COVID-19 vaccination based on real-life public data, in terms of protection against deaths being around 72%, although it appears to be slightly lower than could be expected from clinical trial results. This suggests the absence of indirect protection for non-vaccinated individuals. Results also suggests that vaccination effectiveness against mortality associated with the B.1.1.7 variant is high but slightly lower compared with other variants (70% and 78%, respectively). Finally, this analysis confirms the role of mobility reduction, both within and between countries, as an effective way to reduce COVID-19 mortality and supports the possibility of seasonal variations in COVID-19 incidence.

## Data Availability

Authors confirm that the data used in this study are publicly available and all data sources were listed within the manuscript.

## Acknowledgements

None.

